# Children with Critical Illness Carry Risk Variants Despite Non-Diagnostic Whole Exome Sequencing

**DOI:** 10.1101/2022.05.01.22274445

**Authors:** Joshua E. Motelow, Natalie C. Lippa, Joseph Hostyk, Evin Feldman, Matthew Nelligan, Zhong Ren, Anna Alkelai, Joshua D. Milner, Ali G. Gharavi, Yingying Tang, David B. Goldstein, Steven G. Kernie

## Abstract

Rapid genetic sequencing is an established and important clinical tool for management of pediatric critical illness. The burden of risk variants in children with critical illness but a non-diagnostic exome has not been explored. This was a retrospective case-control analysis of research whole exome sequencing data that first underwent a diagnostic pipeline to assess the association of rare loss-of-function variants with critical illness in children with diagnostic and non-diagnostic whole exome sequencing including those with virally mediated respiratory failure. Using a gene-based collapsing approach, the odds of a child with critical illness carrying rare loss-of-function variants were compared to controls. A subset of children with virally mediated respiratory failure was also compared to controls. Cases were drawn from the general pediatric ward and pediatric intensive care unit (PICU) at Morgan Stanley Children’s Hospital of NewYork-Presbyterian (MSCH) – Columbia University Irving Medical Center (CUIMC) and from the Office of the Chief Medical Examiner (OCME) of New York City. Of the 285 enrolled patients, 228 (80.0%) did not receive a diagnosis from WES. After quality control filtering and geographic ancestry matching, an analysis of 232 children with critical illness compared to 5,322 healthy and unrelated controls determined cases to harbor more predicted loss-of-function (pLOFs) in genes with a LOEUF score ≤ 0.680 (*p*-value = 1.0 × 10^−5^). After quality control and geographic ancestry matching, a subset of 176 children without a genetic diagnosis compared to 5,180 controls harbored pLOFs in genes without a disease association (OR 1.7, CI [1.2 – 2.3], FDR adjusted p-value = 4.4 × 10^−3^) but not in genes with a disease association (OR 1.2, CI [0.8 – 1.7], FDR adjusted p-value = 0.40). This enrichment primarily existed among ultra-rare variants not found in public data sets. Among a subset of 25 previously healthy children with virally mediated respiratory failure compared to 2,973 controls after quality control and geographic ancestry matching, cases harbored more variants than controls in genes without a disease association at the same LOEUF threshold ≤ 0.680 (OR 2.8, CI [1.2 – 7.2], FDR adjusted p-value = 0.026) but not in genes with a disease association (OR 0.7, CI [0.2 – 2.2], FDR adjusted p-value = 0.84). Finally, children with critical illness for whom whole exome sequencing data from both biological parents were available, we found an enrichment of *de novo* pLOF variants in genes without a disease association among 114 children without a genetic diagnosis (unadjusted *p*-value < 0.05) but not among 46 children with a genetic diagnosis. Children with critical illness and non-diagnostic whole exome sequencing may still carry risk variants for their clinical presentation in genes not previously associated with disease.

## Introduction

Children with monogenic or chromosomal abnormalities account for 16% of pediatric admissions and are at increased risk for death in pediatric intensives care units (PICUs).^1-4^ Next generation sequencing (NGS) technology, which includes whole exome sequencing (ES) and whole genome sequencing (GS), achieves a diagnosis in 24%-52% of children with critical illness and frequently changes management.5-12 Among patients with a non-diagnostic ES (ndES), an explanatory variant may be unreported because the variant (1) was undetected by ES, (2) was detected by ES in a gene linked to the patient’s phenotype but it was not determined to be causative or (3) was detected by ES in a gene of unknown significance among other potential reasons.^13,14^

Bronchiolitis refers to a viral infection of the lower respiratory tract.^15^ Nearly every child will be exposed to a virus that causes bronchiolitis by two years of age.^16^ Of children admitted to the hospital for bronchiolitis, ∼9% require ventilatory support and death occurs in less than 0.1%.^17,18^ Although risk factors for hospital admission have been identified including prematurity, most children admitted for bronchiolitis are born at full term without risk factors.^15,18-20^ Genetic risk factors have been proposed but a definitive answer has remained elusive.^21-25^

We hypothesized that children with critical illness and ndES carry deleterious variants in genes without a disease association. These variants are likely risk factors for the child’s illness but do not yield a diagnosis because the genes are not currently associated with the illness. If such a relationship existed, larger studies would be necessary to uncover the genes for which dysfunction is a risk factor for pediatric critical illness including among previously healthy children. We found that children with critical illness carry more loss-of-function variants than controls. Among children without a genetic diagnosis, this signal was strongest among ultra-rare (absent in gnomAD and ExAC) variants in genes without a disease association. We asked the same question of children with virally mediated respiratory failure and found a similar pattern. Finally, we found a larger than expected number of *de novo* loss-of-function variants in genes without a disease association among children without a genetic diagnosis but not with a genetic diagnosis.

## Methods

### Study Population

Children with critical illness were enrolled through two arms: 1) prospective enrollment into research whole exome sequencing (ES) protocols at NewYork-Presbyterian Morgan Stanley Children’s Hospital/Columbia University Irving Medical Center (MSCH/CUIMC, “MSCH cohort”, n = 267) and 2) deceased pediatric cases from the Office of the Chief Medical Examiner (OCME) of New York City (“OCME cohort”, n = 18). Critical illness in the MSCH cohort was defined by either admission to the pediatric intensive care unit (PICU) at MSCH/CUIMC or respiratory failure from a common virus requiring non-invasive positive pressure. Data from the OCME were collected from 2003 to 2016 found using a keyword search for viral bronchiolitis in the immediate cause of death. Of the 24 cases identified, 18 had adequate samples for ES (Table 1, eTable 1). We defined a “combined cohort” by merging the MSCH cohort and the OCME cohort (Table 1, n = 285). A “respiratory failure cohort” was created combining previously healthy probands from the MSCH cohort with respiratory failure secondary to a common virus (n = 22, eTable 2) and (2) probands from the OCME cohort with no phenotypic data suggestive of chronic illness or alternative etiology prior to death (n = 14, eTable 1).

**Table 1:** Summary of Study Participants

|  | Characteristic | No. (%) |
| --- | --- | --- |
| **Sex** |  |  |
|  | **Female** | **137 (48)** |
|  | **Male** | **148 (52)** |
| **Enrollment location** |  |  |
|  | **Clinic** | **131 (46)** |
|  | **PICU** | **136 (48)** |
|  | **OCME** | **18 (6)** |
| **Diagnostic Status** |  |  |
|  | **Resolved** | **46 (16)** |
|  | **Partially Resolved** | **11 (4)** |
|  | **Unresolved** | **228 (80)** |
| **Geographic Ancestry** |  |  |
|  | **Admixed** | **37 (13)** |
|  | **African** | **87 (31)** |
|  | **East Asian** | **12 (4)** |
|  | **European** | **42 (15)** |
|  | **Latino** | **92 (32)** |
|  | **Middle Eastern** | **1 (0)** |
|  | **South Asian** | **14 (5)** |
| **Trios** |  |  |
|  | **Resolved** | **46** |
|  | **Unresolved** | **114** |

### Controls

Controls were drawn from unrelated individuals with data stored at the Institute for Genomic Medicine (IGM) at CUIMC enrolled as either “controls” (n = 3,358) or “healthy family members” (n = 6,632) across multiple studies.

### Next Generation Sequencing, Variant Calling, and Quality Control

Sequencing of cases and control individuals was performed at or transferred to the IGM. Case data are all ES. Controls were a mixture of ES (n = 9,353) and whole genome sequencing (GS, n = 637). Established protocols have been used at the IGM to record to generate ES or GS data for more than 120,000 individuals (eMethods).^26-28^ Variants were annotated with IGM’s in-house analysis tool for annotated variants (ATAV) platform.^29^

### Diagnostic Analysis

The diagnostic analysis and outcomes of the MSCH cohort are a subset of the cohort reported by Lippa and colleagues.^30^ Briefly, trio or proband-only ES analysis was completed. Candidate genotypes were reviewed with the research team and clinical team to reach a consensus interpretation. All variants deemed highly likely to be causative were confirmed in a Clinical Laboratory Improvement Amendments (CLIA) certified laboratory prior to being returned to the patients and their providers.

Probands that harbored a genetic finding which explained all or part of their phenotypes were deemed “resolved” (Table 1, n = 57). The remainder were considered the “unresolved cohort” (n = 228). The ES from these probands were considered non-diagnostic ES (ndES). Samples recruited from the OCME cohort underwent a modified diagnostic pipeline due to the limited phenotypic information (eTable 1). No explanatory variants were identified in this cohort, and all were considered “unresolved”.

### Clustering

We matched cases and controls by geographic ancestry using clusters as previously described (eFigure 1).^31-33^ Principal Component Analysis (PCA) for dimensionality reduction was performed on a set of predefined variants to capture population structure.^34^ Clusters were identified using the Louvain method of community detection using the first six principal components (PCs) as input which reflect the geographic ancestry of the samples.^31,35^

### Gene-Based Collapsing

Collapsing refers to an analysis where variants satisfying specific criteria (qualifying variants [QVs]) are considered equivalent and a statistical association is determined between harboring a QV in a gene or gene-set and case/control status. QV specification then determines the collapsing model (eTable 3).^31,33,36,37^ For each gene and individual within each cluster, an indicator variable (1/0 states) was assigned based on the presence of at least one QV in the gene (state 1) or no qualifying variants in that gene (state 0) to create a gene-by-individual matrix for each cluster. From the collapsing matrices of each cluster, we extracted the number of cases/controls with and without a QV per gene and tested for an enrichment of qualifying variants in the case or control group for each gene.^31,33,38-40^

### Loss-of-Function Analysis

We assessed the optimal gene intolerance threshold associating case/control status and QV carrier status (Figure 1).^14^ In the Rare pLOF model (eTable 3) in the combined cohort, 8,702 genes with 1,860 unique loss-of-function observed/expected upper bound fraction (LOEUF) scores harbored QVs.^41^ These genes were ordered based on their LOEUF scores from most to least intolerant to loss-of-function variation and gene-sets were created for genes less than or equal to each LOEUF threshold. For each gene-set, we calculated a *p*-value using the CMH signifying the statistical association of case/control status and pLOF carrier status. Empiric *p*-values were determined by 100,000 permutations of the CMH *p*-values. We used a Bonferroni correction for the 1,860 comparisons to assess significance (0.05 / 1,860).

**Figure 1:**
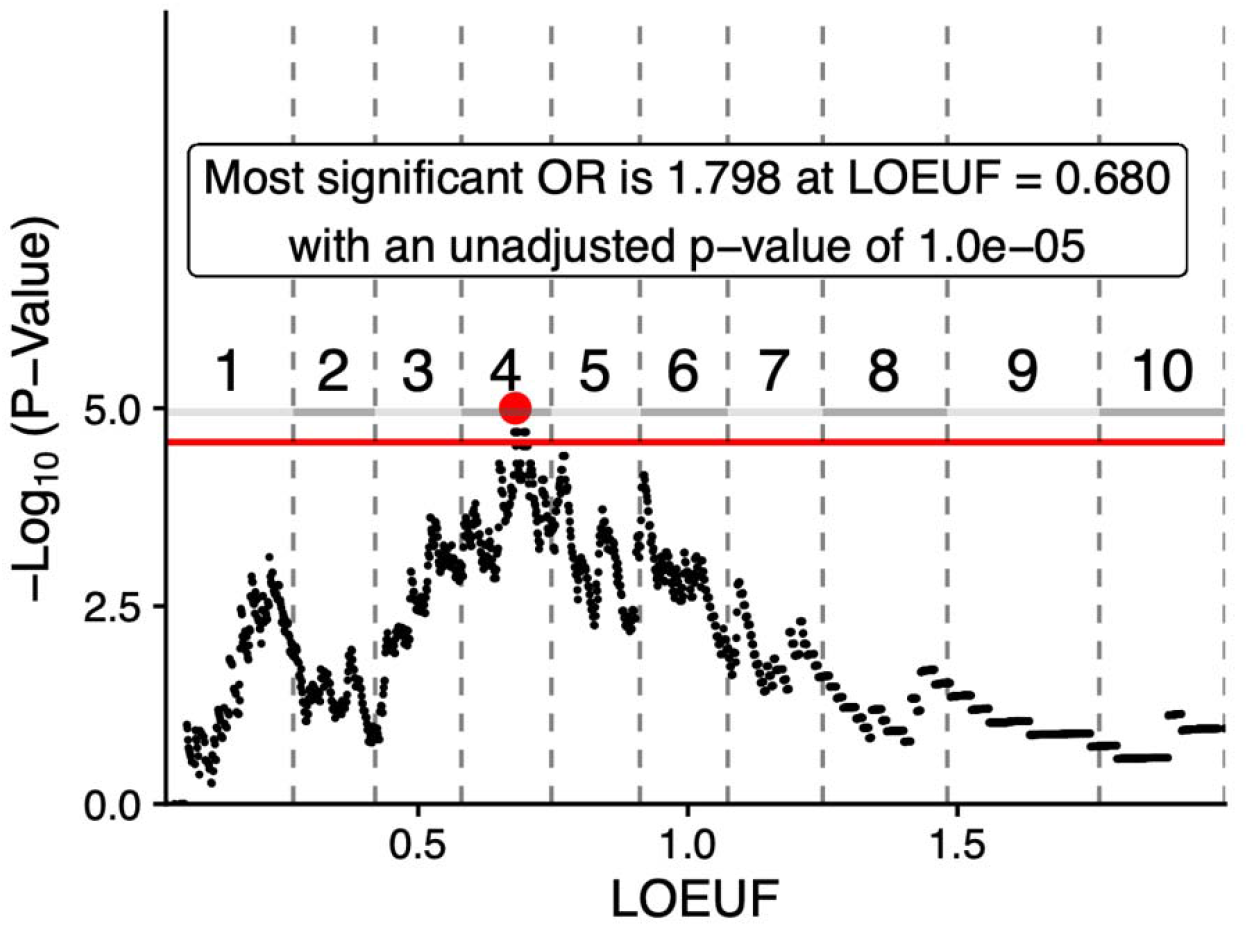
Children with critical illness harbor more loss-of-function variants than controls. We used an unbiased approach to explore the hypothesis that children with critical illness (231 cases and 5,322 controls) were enriched with pLOF variants and that this enrichment was related to genic intolerance. We examined 1,860 unique gene sets created by taking genes equal to or less than each of the 1,860 unique LOEUF scores found in genes carrying pLOF variants in our combined case/control cohort. Gene-sets to the left of the plot are more intolerant to loss-of-function variation sub-sets of gene-sets to the right of the plot. For each gene-set, enrichment was evaluated with the use of the exact two-sided Cochran-Mantel-Haenszel (CMH) statistic that evaluated case–control status regarding the presence or absence of a loss-of-function variant in each gene grouping producing a pooled odds ratio and *p-*values. Empiric p-values were determined by 100,000 permutations. The horizontal redline represents significance using Bonferroni correction (red line, 0.05 / 1860). The negative log of the permuted p-value at each LOEUF threshold is shown on the y axis. The minimum empirically derived *p*-value, which indicates the most significant enrichment across all LOEUF thresholds, occurs at a LOEUF ≤ 0.680. The 1 to 10 labels indicate the intolerance decile based on LOEUF scores with “1” representing the 10% most intolerant genes and “10” representing the 10% least intolerant genes.

### Gene-Set Collapsing

We tested associations of case/control status with QV status in the following gene-sets (eTable 4). Genes with a confirmed phenotype in Online Mendelian Inheritance in Man (OMIM) on 02/16/2021 (3,964 genes).^42^ Of these, 1,931 harbored a variant in a non-synonymous model. Of the genes without a confirmed phenotype in OMIM, 6,821 harbored a variant in a non-synonymous model. We included a gene panel for primary immunodeficiencies from Invitae and a gene panel associated with susceptibility to life-threatening viral illness.^43^

### *De Novo* Mutation Calling, Filtering and Analysis

We obtained *de novo* variants for probands with both biological parents enrolled. To test the null hypothesis that our cohort contained an expected number of *de novo* variants in genes without a disease association, we utilized denovolyzeR.^44-46^ denovolyzeR is an R package which analyzes the rate of *de novo* variants in a dataset and compares it to an expected variant rate to assess enrichment.

## Results

### Diagnostic Outcomes and Phenotypes

This analysis included 285 children with critical illness (Table 1) enrolled either from MSCH/CUIMC or the OCME aged equal to or less than 18 years at enrollment (median = 4.1 years, range 0 – 18.9 years). 57 of 285 achieved a full or partial diagnosis (20.0%) (eTable 5). The phenotypes enrolled were heterogenous (eTable 6).

### Clustering and Collapsing

In the combined, unresolved, and respiratory failure cohorts, 231 cases and 5,322 controls, 176 cases and 5,180 controls, and 25 cases and 2,973 controls were analyzed, respectively, after QC and ancestry matching (eTable 7) for cluster-based, rare-variant collapsing association analyses. For each cohort, two models were tested with pLOF variants (eTable 3). The ultra-rare pLOF model included only variants absent from public datasets while a more inclusive Flex pLOF model allowed variants with a minor allele frequency (MAF) < 0.1%. No model showed a single gene association with study-wide significance (eFigures 2 – 10, eTables 8 - 16). The clusters included in the analysis encompassed cases and controls of European, Latino, and African geographic descent (eFigure 1). The rare synonymous variant association analysis functioned as a control and the low genomic-inflation factor indicated adequate sub-structure matching (eFigures 2 - 4, eTables 8 – 10).

### pLOF Burden in Children with Critical Illness

We assessed whether children with critical illness carry more pLOF variants than controls and whether this relationship is mediated by genic intolerance (Figure 1).^14^ We identified 8,702 unique genes harboring a rare pLOF variant in either a case or control (MAF < 0.1%, “Flex pLOF” mode, eFigure 8, eTable 3, 14), which comprised 1,860 unique LOEUF scores.^41^ The LOEUF score quantifies a gene’s intolerance to loss-of-function variation. We created 1,860 gene-sets comprised of genes less than or equal to the unique LOEUF scores and ordered the gene-sets from most to least intolerant. We assessed association of children with critical illness and carrier status in each gene-set. A statistically significant association between case/control status and pLOF variants exists, and the LOEUF threshold with the most significant association was 0.680 (OR = 1.8, unadjusted *p*-value by permutation = 1.0 × 10^−5^).

We asked three questions: are the variants present in public databases? Are the genes harboring the variants associated with diseases? Is the association driven by children with a genetic diagnosis? We considered genes with a LOEUF score ≤ 0.680 (“thresholded genes”), which was the threshold at which pLOF carrier status and case/control status was strongest (Figure 1). In the combined cohort (Figure 2A, eTable 17), the relationship between pLOF variants and children with critical illness exists in both genes with a disease association (CMH, OR = 1.6, CI [1.2 2.2], *adj*.*p* = 6.4 × 10^−3^) and without (CMH, OR = 1.5, CI [1.1 2.0] *adj*.*p* = 8.6 × 10^−3^).^42^ The association was only significant for variants absent from public datasets in genes (CMH, OR = 1.9, CI [1.3 2.6], *adj*.*p* = 2.3 × 10^−3^) and without (CMH, OR = 1., CI [1.1 2.0], 5, *adj*.*p* = 0.016). Carrying variants which were rare (MAF < 0.1%) but still present in public datasets was not associated with case-control status regardless of gene-disease associations. This supports the hypothesis that rare diseases (with pediatric critical illness representing either an unusual illness or an unusually severe presentation of a common illness) are driven by variants absent in public datasets.^40^

**Figure 2:**
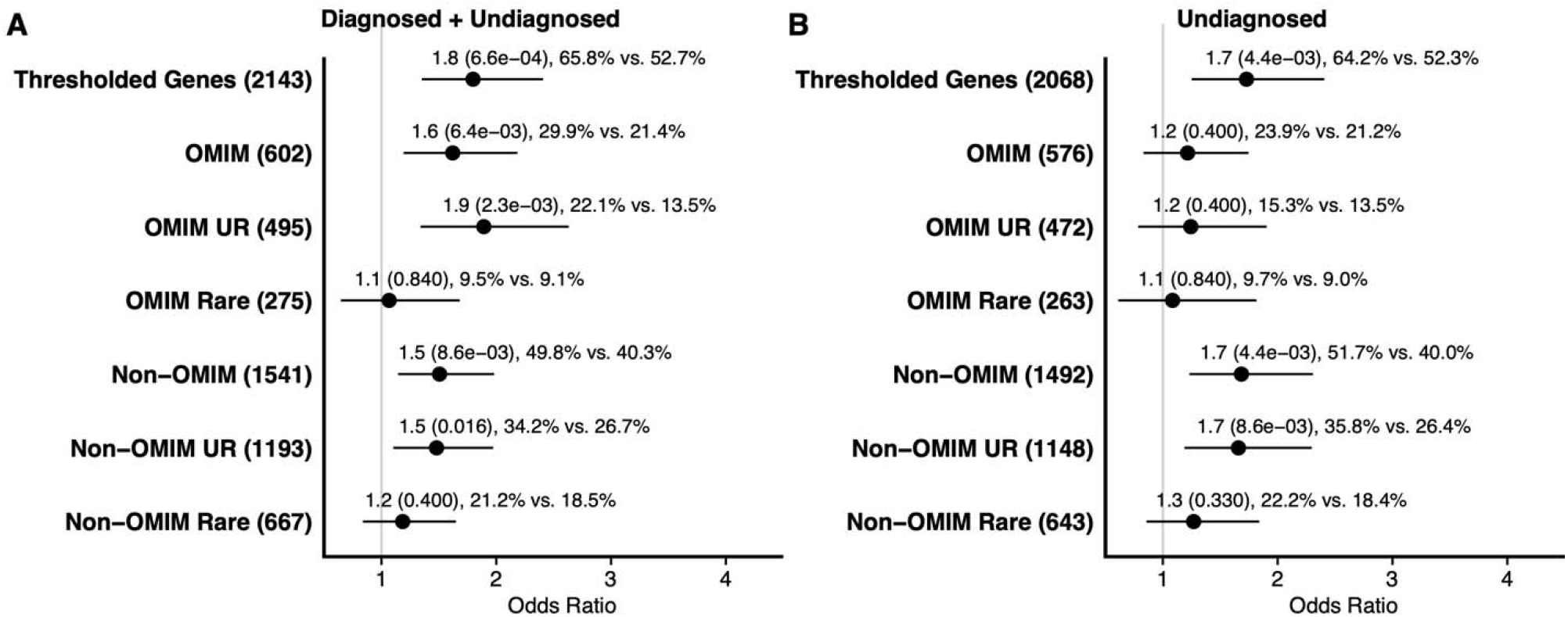
Gene-Set Enrichment Analysis Juxtaposing the Combined Cohort against the Unresolved Cohort. Gene-set burden analysis to understand the source of predicted loss-of-function (pLOF) signal associated with children with critical illness. Genes with a LOEUF score ≤ 0.680 (“Thresholded Genes”) harbor the most significant association with children with critical illness among all the LOEUF scores tested (Figure 1). For the (A) combined cohort (231 cases and 5,322 controls) and (B) unresolved (i.e., no genetic diagnosis) cohort (176 cases and 5,180 controls). Variants are divided into those in disease genes (“OMIM”) and genes without a disease association (“Non-OMIM”), ultra-rare (“UR” [absent from external control datasets]) and rare (“Rare” [minor allele frequency < 0.1% but present in external control datasets]). Variants are all high-quality pLOF (see Methods). Pooled odds ratio, 95% confidence intervals and FDR corrected *p-*value were generated from the exact two-sided Cochran-Mantel-Haenszel (CMH) test. X-axis displays the odds ratio and confidence intervals.

We removed children carrying variants partially or fully explaining their phenotypes to separately assess the exomes children without a genetic diagnosis (unresolved cohort, Figure 2B, eTable 18). The association of carrying a pLOF variant in genes with a disease association and case/control status was not significant (CMH, OR = 1.2, CI [0.8 1.7], *adj*.*p* = 0.40). Interestingly, the association of carrying a pLOF variant in genes without a disease association remained significant (CMH, OR = 1.7, CI [1.2 2.3], *adj*.*p* = 4.4 × 10^−3^). This supports the hypothesis that children with ndES carry ultra-rare risk variants in genes that are not yet associated with a disease phenotype. As in the combined cohort, this relationship extended to variants absent from public databases (CMH, OR = 1.7, CI [2.2 2.3], *adj*.*p* = 8.6 × 10^−3^) but not rare variants (MAF < 0.1%) present in public databases (CMH, OR = 1.3, CI [0.9 1.8], *adj*.*p* = 0.33).

### Virally Mediated Respiratory Failure may be Associated with Genetic Risk

We assessed whether harboring an ultra-rare pLOF variant in a “thresholded gene” might be a risk factor for virally mediated respiratory failure (Figure 3, eTable 19). There was no significant association between case-control designation and carrier status of pLOF variants in genes with a disease association (CMH, OR = 0.7, CI [0.2 2.2], *adj*.*p* = 0.84). Interestingly, for genes without a disease association, the association was significant (CMH, OR = 2.8, CI [1.2 7.2], *adj*.*p* = 0.026) and was strengthened when considering only variants absent from public databases (CMH, OR = 3.3, CI [1.4 8.1], *adj*.*p* = 9.3 × 10^−3^). We found no association among genes implicated in primary immunodeficiencies or virally mediated respiratory failure (eTable 20).^18,21,22,25,43,47-52^

**Figure 3:**
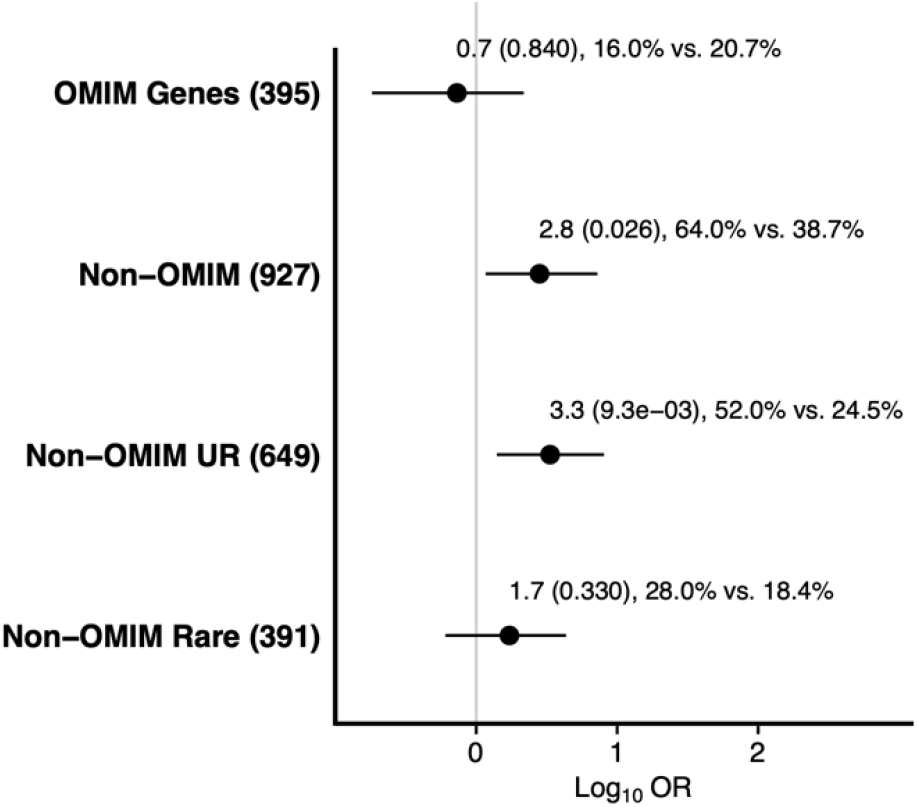
Children with viral respiratory failure harbor ultra-rare loss-of-function genetic risk factors. Gene-set burden analysis to understand the source of predicted loss-of-function (pLOF) signal associated with children with viral respiratory failure. Genes with a LOEUF score ≤ 0.680 harbor the most significant association with children with critical illness among all the LOEUF scores tested (Figure 1). Data include 25 cases compared to 2,973 controls. Genes associated with a disease (“OMIM”) and not associated with disease (“Non-OMIM”) were investigated. Variants are divided into ultra-rare (“UR” [absent from external control datasets]) and rare (“Rare” [minor allele frequency < 0.1% but present in external control datasets]). Variants are all high-quality pLOF (see Methods). Odds ratio, 95% confidence intervals and FDR corrected *p-*value were generated from the Fisher’s exact test. X-axis displays the logarithm of the odds ratio and confidence intervals.

Our respiratory failure cohort sample size was inadequate to assess gene-specific associations (eFigures 7, 10 eTables 13, 16). To identify candidate genes associated with risk of viral mediated respiratory failure, we highlighted 12 “thresholded genes” without a disease association harboring ultra-rare pLOF variants in cases but not controls (eTable 21). Candidate genes include *CTTN* (MIM: 164765) which is implicated in lung disease among patients of African descent,^53^ *RSBN1L* (MIM: N/A) which is upregulated in severe COVID-19,^54^ and *MKL2* (MIM: 609463), a gene closely related to *MKL1* (MIM: 606078), which is implicated immunodeficiency 66 (MIM: 618847).^55-57^ *RASGRP4* (MIM: 607320) regulates dendritic and mast cell function.^58,59^ No genes harbored ultra-rare pLOFs in multiple probands in the respiratory failure cohort suggesting genetic heterogeneity.

### De Novo Enrichment in Children with Critical Illness

Among the 57 resolved or partially resolved probands, 46 were trios (i.e., ES available from the proband and both biological parents) and among the 227 unresolved probands, 114 were trios (Table 1). Among the 160 total trios, we assessed enrichment of *de novo* variants in genes without a disease association (Table 2, eTable 8).^44,46^ *De novo* pLOF variants were enriched in unresolved probands (*p*-value = 0.040) but not in fully or partially resolved probands (*p*-value = 0.50). These data further the hypothesis that despite a ndES, probands in the PICU likely carry pLOF risk variants for their phenotype.

**Table 2:** Enrichment of *de novo* variants in children with critical illness in genes without a disease association The resolved trios (n = 46) and unresolved trios (n = 14) were examined. We examined enrichment among genes without a disease association. Enrichment is limited to loss-of-function variants in the unresolved cohort. P-values were not adjusted for multiple comparisons. *Denovolyzer* was utilized to assess for *de novo* enrichment against the predicted null distribution. Syn = synonymous, Mis = missense, LoF = loss-of-function, Prot = missense + loss-of-function, All = Syn + Mis + LoF.

| *Cohort* | *Variant Class* | *Observed* | *Expected* | *Enrichment* | *P-Value* |
| --- | --- | --- | --- | --- | --- |
| Resolved | Syn | 6 | 8.80 | 0.68 | 8.71e-01 |
| Resolved | Mis | 9 | 19.70 | 0.46 | 9.98e-01 |
| Resolved | LoF | 3 | 2.70 | 1.12 | 5.04e-01 |
| Resolved | Prot | 12 | 22.40 | 0.54 | 9.94e-01 |
| Resolved | All | 18 | 31.20 | 0.58 | 9.96e-01 |
| Unresolved | Syn | 10 | 21.80 | 0.46 | 9.98e-01 |
| Unresolved | Mis | 42 | 48.90 | 0.86 | 8.57e-01 |
| Unresolved | LoF | 12 | 6.70 | 1.80 | 3.97e-02 |
| Unresolved | Prot | 54 | 55.60 | 0.97 | 6.03e-01 |
| Unresolved | All | 64 | 77.40 | 0.83 | 9.46e-01 |

## Discussion

In this, the first study to our knowledge to examine the exomes of critically ill children with non-diagnostic ES, we found that a non-diagnostic exome may identify a disease-risk variant in genes not associated with a disease. The diagnostic rate of 20.0% (Table 1) was lower than that of other PICU cohorts, but our inclusion criteria allowed enrollment whether the proband was suspected to have a genetic condition or not. Therefore, the diagnostic yield is lower than an approach meant to identify patients with a high pre-test probability.^10,60^ The prevalence of seizures and respiratory failure Human Phenotype Ontology terms (eTable 6), though common PICU admission diagnoses,^17,61^ also reflects recruitment strategies.

We found that carrying a pLOF variant in genes with a LOEUF score ≤ 0.680 was significantly associated with cases compared to controls (Figure 1). The importance of this LOEUF threshold is challenging to interpret. A cohort of stillbirth cases undergoing a similar analysis found a peak LOEUF threshold to be 0.239.^14^ Stillbirth is a more severe phenotype and therefore we might expect the peak LOEUF threshold to capture more intolerant genes than children who have survived the neonatal period. A future analysis to assess whether the peak LOEUF threshold is related to disease severity could include a collection of phenotypically homogenous cohorts with a range of disease severities.

Among the combined cohort, genes with and without a disease association harbored pLOFs (Figure 2A). Among children with critical illness without a genetic diagnosis, the association with disease-associated genes was lost while the association with genes without a disease association remained (Figure 2B). This implies that critically ill children with non-diagnostic ES may still carry variants which are a risk factor for their phenotype, but no diagnosis is made because the gene-disease association is not understood. Comparing the rate of carrier status of pLOF variants in genes with a LOEUF score ≤ 0.680 among unresolved cases (51.7% in cases vs. 40.0% in controls), we can estimate that ∼10% of children in the PICU without a genetic diagnosis may ultimately be diagnosed with improved gene-disease association discovery for pediatric critical illness. These risk variants are likely to be absent from public databases (Figures 2 and 3).

Despite a small sample size of children with virally mediated respiratory failure, we found a significant burden of pLOFs in genes without a disease association with a LOEUF ≤ 0.680 (Figure 3). We highlighted three candidate causative genes (*CTTN, RSBN1L, MKL2, RASGRP4*). No single gene harbored variants in more than one case suggesting genetic risk is heterogenous. Though small, these data indicate that a larger study could yield statical evidence of gene-disease association.^33^

Finally, we examined the enrichment of *de novo* variants (Table 2). Among children with critical illness without a diagnosis, we found more *de novo* loss-of-function variants than expected in genes without a disease association. These variants may, in fact, be determined to be causative as gene-phenotype connections are discovered.

### Limitations

Limitations of this study include the small sample size of the cohort, heterogeneity of phenotypes, absence of standard additional genetic testing such as chromosomal microarray and the absence of full phenotyping in OCME patients. It is our hope that increasing sample sizes of children with critical illness will allow the discovery of gene-disease associations for specific phenotypes.

## Conclusions

We found that children with critical illness harbor more loss-of-function variants than controls. Among children without a genetic diagnosis, these variants were ultra-rare and resided in genes without a disease association. We found this association to be true among a subset of patients with virally mediated respiratory failure despite a small sample size. Finally, among children without a genetic diagnosis, we found more than expected *de novo* pLOF variants in genes without a disease association. Our data indicate that children with critical illness may harbor ultra-rare risk variants for their phenotypes, which are detected by ES even in the absence of a genetic diagnosis.

## Data Availability

All data produced in the present study are available upon reasonable request to the authors

## Acknowledgements

We thank Katherine Schlosser Metitiri, MD for her assistance in determining PICU admission status. J.E.M. is supported by a Thrasher Early Career Research Award. A.G.G. has received research funding from the Renal Research Institute and Natera and has served as a consultant for Goldfinch Bio, Actio Biosciences, Travere and Novartis. D.B.G. holds equity in Q State Bioscience and is a founder and holds equity in Actio Biosciences, Inc and Praxis Therapeutics; holds equity in Apostle Inc., and serves as a consultant to AstraZeneca, Gilead Sciences, GoldFinch Bio and Gossamer Bio and as full time CEO of Actio Biosciences.

