## Supplemental Tables Figures and Methods for "Children with Critical Illness Carry Risk Variants Despite Non-Diagnostic Whole Exome Sequencing"

### Title

Supplemental Tables

#### Supplementary Tables

| *Immediate Cause of Death* | *Ancestry Matched in Combined Cohort* | *Respiratory Failure Cohort* | *Adequate Ancestry Match in Respiratory Failure Cohort* |
|---|---|---|---|
| Viral Bronchiolitis | X | X | X |
| Bronchiolitis and Interstitial Pneumonia, (Probable Viral) | X | X | X |
| Acute Bronchiolitis and Early Bronchopneumonia due to Prematurity | X | | X |
| Bronchiolitis and Interstitial Pneumonia, (Probable Viral) | X | X | X |
| Bronchiolitis and Interstitial, Viral - Type, Pneumonia | X | X | X |
| Acute bronchopneumonia and bronchiolitis complicating interstitial pneumonia, (probable viral etiology) | X | X | X |
| Upper Respiratory Tract Infection, Probably Viral, Complicated By Bronchitis And Bronchiolitis | X | X | X |
| Klebsiella Bronchopneumonia Complicating Bronchiolitis | X | X | X |

| | | | |
|---|---|---|---|
| Chronic laryngitis, tracheitis, bronchitis and bronchiolitis due to probable viral infection. Chronic bronchial asthma and Anamolous R coronary artery origin at left sinus w/ acutely angled takeoff | X | | X |
| Respiratory syncytial virus bronchiolitis | X | X | X |
| Acute viral bronchiolitis with bronchopneumoni a | X | X | |
| Adenoviral bronchiolitis | | X | |
| Chronic bronchiolitis | X | | X |
| Complications of Parainfluenza 3 viral infection including myocarditis, | | | |
| Acute Bronchopneumoni a complicating Tracheitis, Bronchitis and Bronchiolitis of Probable Viral Etiology | | X | |
| Bronchiolitis of probable viral origin | X | X | X |
| Acute Bronchiolitis | | X | |
| Bronchiolitis and Pneumonitis | X | X | X |

*eTable 1: Probands recruited from the Office of the Chief Medical Examiner of New York City (OCME)*

Summary of limited clinical information available from the OCME and inclusion in analysis. All subjects were aged less than 5 years old.

| *Virus* | *Respiratory Support* | *Analyzed Cluster* |
| --- | --- | --- |
| respiratory syncytial virus rna | CMV | x |
| respiratory syncytial virus rna | CMV | |
| human rhinovirus/enterovirus rna | BPAP | x |
| human rhinovirus/enterovirus rna, influenza? Need to double check chart | BPAP | x |
| respiratory syncytial virus rna | CMV | |
| human rhinovirus/enterovirus rna, respiratory syncytial virus rna | BPAP | x |
| respiratory syncytial virus rna | CPAP | |
| human rhinovirus/enterovirus rna | BPAP | x |
| human rhinovirus/enterovirus rna | CMV | x |
| adenovirus, human metapneumovirus rna | HFOV | |
| human rhinovirus/enterovirus rna | BPAP | x |
| human rhinovirus/enterovirus rna | CPAP | x |
| human rhinovirus/enterovirus rna | HFOV | x |
| human rhinovirus/enterovirus rna | CMV | |
| human rhinovirus/enterovirus rna | BPAP | x |
| human rhinovirus/enterovirus rna | CMV | |
| human rhinovirus/enterovirus rna, respiratory syncytial virus rna | BPAP | x |
| respiratory syncytial virus rna | CMV | |
| respiratory syncytial virus rna | BPAP | |
| human rhinovirus/enterovirus rna | CPAP | x |
| respiratory syncytial virus rna | BPAP | x |
| respiratory syncytial virus rna | CMV | x |

*eTable 2: Summary of MSCH Respiratory Failure Cohort.*

All subjects aged less than 5 years old. BPAP = bilevel positive airway pressur, CPAP = continuous positive airway pressure, CMV = conventional mechanical ventilation, HFOV = high frequency oscillatory ventilation

| *Model Name* | *Included Effects* | *GnomAD Exome Max AF Population Specific* | *GnomAD Genome Max AF Global* | *Cohorts Analyzed Independently with Model* |
|---|---|---|---|---|
| Ultra-Rare Synonymous | Synonymous | 0 | 0 | Combined, Unresolved, Viral Respiratory Failure |
| Ultra-Rare pLOF | pLOF | 0 | 0 | Combined, Unresolved, Viral Respiratory Failure |
| Flex pLOF | pLOF | 0.001 | 0.001 | Combined, Unresolved, Viral Respiratory Failure |

*eTable 3: Collapsing Model Qualifying Variant Definition*
Summary of collapsing models and qualifying variant definitions. pLOF = predicted loss-of-function effect. pLOF effects include stop gained, frameshift, splice acceptor, and splice donor variants.

In a separate Excel file (large tables):

*eTable 4: Genes with and without Disease Associations in OMIM*

Summary of genes harboring a pLOF in any of non-synonymous models in this study and the genes association with a disease in OMIM.

*eTable 5: Summary of Diagnostic Variants*

Table summarizing the diagnostic variants of the children whose phenotypes are fully (n = 46) or partially (n = 11) by the genetic finding.

| *HPO Label* | *HPO ID* | *Count* |
| --- | --- | --- |
| Respiratory failure | HP:0002878 | 110 |
| Seizure | HP:0001250 | 108 |
| Severe viral infection | HP:0031691 | 76 |
| Fever | HP:0001945 | 69 |
| Respiratory distress | HP:0002098 | 69 |
| Hypoxemia | HP:0012418 | 50 |
| Tube feeding | HP:0033454 | 49 |
| Respiratory failure requiring assisted ventilation | HP:0004887 | 47 |
| Feeding difficulties | HP:0011968 | 47 |
| Cough | HP:0012735 | 45 |

*eTable 6: Top HPO terms*

| *Cohort* | *Initial Cohort* | *After QC* | *After Relatedness Check* | *Cases Included in Analyzed Clusters* | *Controls Included in Analyzed Clusters* |
|---|---|---|---|---|---|
| All | 285 | 282 | 280 | 231 | 5322 |
| Unresolved | 229 | 226 | 225 | 156 | 3619 |
| Respiratory Failure | 36 | 36 | 36 | 25 | 2973 |

*eTable 7: Samples Removed during Data Cleaning*

In a separate Excel file (large tables):
*eTable 8: Top 50 Genes in Synonymous Model in Combined Cohort*
*eTable 9: Top 50 Genes in Synonymous Model in Unresolved Cohort*
*eTable 10: Top 50 Genes in Synonymous Model in Respiratory Failure Cohort*
*eTable 11: Top 50 Genes in pLOF Ultra-Rare Model in Combined Cohort*
*eTable 12: Top 50 Genes in pLOF Ultra-Rare Model in Unresolved Cohort*
*eTable 13: Top 50 Genes in pLOF Ultra-Rare Model in Respiratory Failure Cohort*
*eTable 14: Top 50 Genes in pLOF Flex Model in Combined Cohort*
*eTable 15: Top 50 Genes in pLOF Flex Model in Unresolved Cohort*
*eTable 16: Top 50 Genes in pLOF Flex Model in Respiratory Failure Cohort*
For Tables S4 – S12, summary of top 50 genes in collapsing analysis indicated in the table title. The last two columns indicate gene group membership. O = 3,964 genes with a disease association in OMIM (see eTable 13, eMethods).

*eTable 17: Loss-of-Function Burden in Children with Critical Illness*
Gene set analysis showing the increased burden of loss-of-function variants in children with critical illness among genes with a LOEUF score ≤ 0.680 which was identified as the threshold of peak burden (see Figure 1). Table shows data for Figure 2a (see figure legend for details).

*eTable 18: Loss-of-Function Burden in Children with Critical Illness but without an Identified Causative Genetic Diagnosis*
Gene set analysis showing the increased burden of loss-of-function variants in children with critical illness but without an identified causative genetic diagnosis among genes with a LOEUF score ≤ 0.680 which was identified as the threshold of peak burden (see Figure 1). Table shows data for Figure 2b (see figure legend for details).

*eTable 19: Loss-of-Function Burden in Children with Viral Mediated Respiratory Failure in Genes with and without a Disease Association*
Gene set analysis showing the increased burden of loss-of-function variants in children with respiratory among genes with a LOEUF score ≤ 0.680 which was identified as the threshold of peak burden (see Figure 1). Table shows data for Figure 3 (see figure legend for details).

*eTable 20: Loss-of-Function Burden in Children with Viral Mediated Respiratory Failure in Immunodeficiency Gene Sets*
Gene-set burden analysis to understand the role of genes associated with immunodeficiencies among children with virally mediated respiratory failure. Data include 25 cases compared to 2,973 controls. Gene-set analysis focused on genes drawn from the two sets. "Primary Immunodeficiency" includes 412 genes drawn from the "Invitae Primary Immunodeficiency Panel" and "Viral Pathway" genes which are drawn from genes previously implicated in virally mediated respiratory failure. Variants are rare ("Rare" [minor allele frequency < 0.1%]). Variants are all high-quality pLOF (see Methods). Pooled odds ratio, 95% confidence intervals and FDR corrected *p*-value were generated from the exact two-sided Cochran-Mantel-Haenszel (CMH) test.

| |
|---|
| ADAMTS7 |
| B3GNT5 |
| CTTN |
| FAM118B |
| GRM5 |
| KIAA1211 |
| MKL2 |
| RASGRP4 |
| RSBN1L |
| WIPI1 |
| ZNF746 |
| FEM1B |

*eTable 21: Candidate Genes Proposed to Harbor Risk Variants Associated with Risk for Virally Mediated Respiratory Failure*

Genes with LOEUF score ≤ 0.680 harboring ultra-rare pLOF variants in cases but not controls in the respiratory failure cohort

**Capture kits used in exome samples**

| *Capture Kit* | *Case* | *Ctrl* |
|---|---|---|
| 65MB | 0 | 542 |
| AgilentV4 | 0 | 109 |
| Agilentv5 | 0 | 4 |
| AgilentV5 | 0 | 8 |
| AgilentV5UTR | 9 | 51 |
| AgilentV6 | 0 | 2 |
| IDTERPv1 | 198 | 2285 |
| MedExome | 0 | 5 |
| Roche | 78 | 6923 |
| RocheV2 | 0 | 32 |

*eTable 22: Capture kits used in whole exome sequencing generation*

#### Supplementary Figures

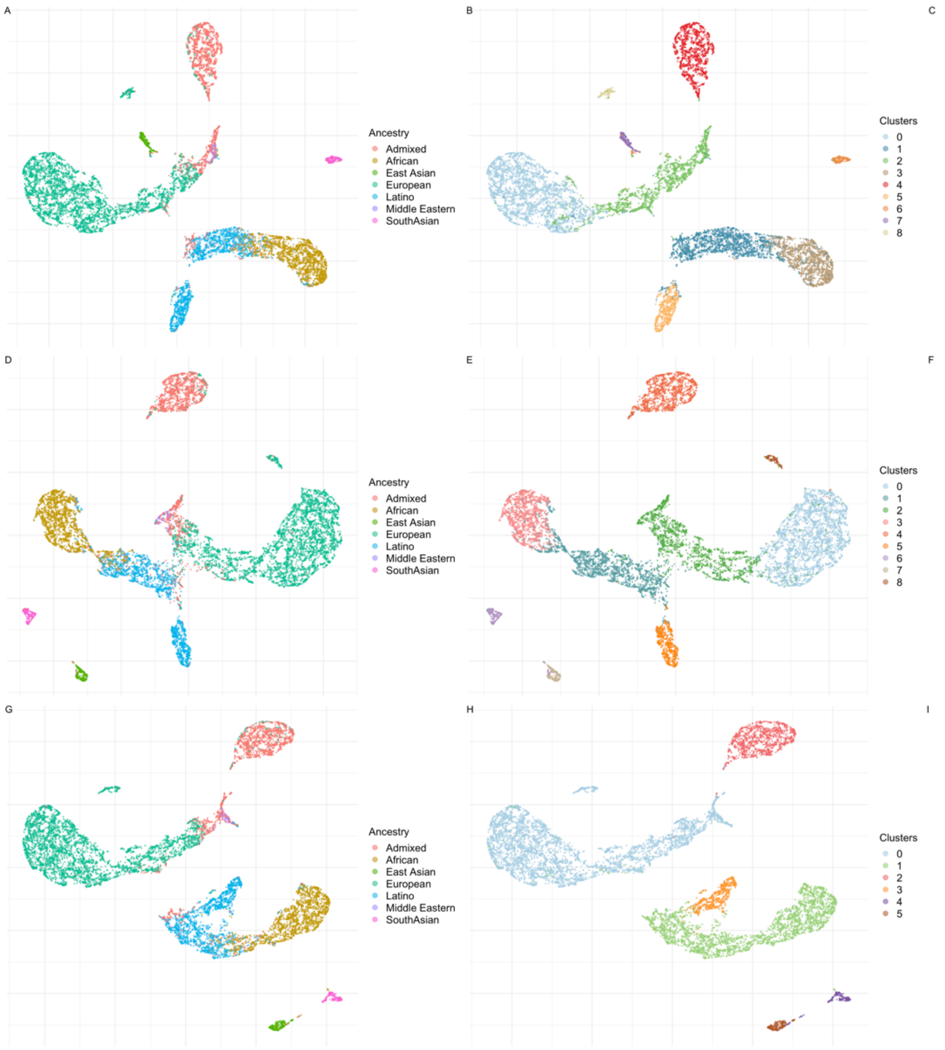

C

| Cluster | Cases | Controls |
|---|---|---|
| 0 | 19 | 2657 |
| 1 | 96 | 1676 |
| 2 | 29 | 1538 |
| 3 | 48 | 1294 |
| 4 | 17 | 1132 |
| 5 | 37 | 606 |
| 6 | 21 | 208 |
| 7 | 10 | 188 |
| 8 | 4 | 129 |

F

| Cluster | Cases | Controls |
|---|---|---|
| 0 | 15 | 2657 |
| 1 | 76 | 1727 |
| 2 | 20 | 1561 |
| 3 | 45 | 1272 |
| 4 | 13 | 1137 |
| 5 | 35 | 620 |
| 6 | 11 | 220 |
| 7 | 7 | 195 |
| 8 | 3 | 130 |

I

| Cluster | Cases | Controls |
|---|---|---|
| 0 | 3 | 4321 |
| 1 | 25 | 2973 |
| 2 | 2 | 1133 |
| 3 | 3 | 609 |
| 4 | 1 | 204 |
| 5 | 2 | 188 |

*eFigure 1: Geographic ancestry and clustering for combined cohort*
UMAP and cluster assignments showing ancestry of case-control cohorts for the combined cohort (A-C), unresolved cohort (D-F), and viral respiratory failure cohort (G-I). Clusters shaded in red were excluded from the analysis.

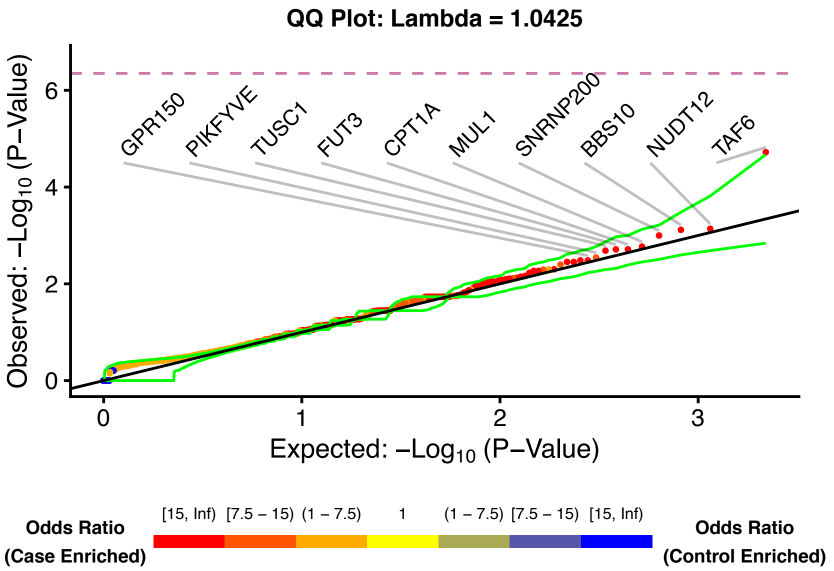

*eFigure 2: Ultra-Rare Synonymous Model Quantile-Quantile Plot for Combined Cohort*
The quantile-quantile plots for the protein-coding genes with at least one case or control carrier of an ultra-rare synonymous variant. All variants are ultra-rare (qualifying variants were defined as a minor allele frequency of less than 0.05% in internal case and control by cluster and absent in external reference cohorts). P-values were generated from the exact two-sided Cochran-Mantel-Haenszel (CMH) test by gene by cluster to indicate a different carrier status of cases in comparison to controls. Study-wide significance $p < 4.5 \times 10^{-7}$ after Bonferroni correction indicated by dashed line (see Collapsing by Gene and Statistical Enrichment. Top ten case enriched genes are labeled. Point coloring determined by CMH odds ratio. The green lines represent the 95% confidence interval.

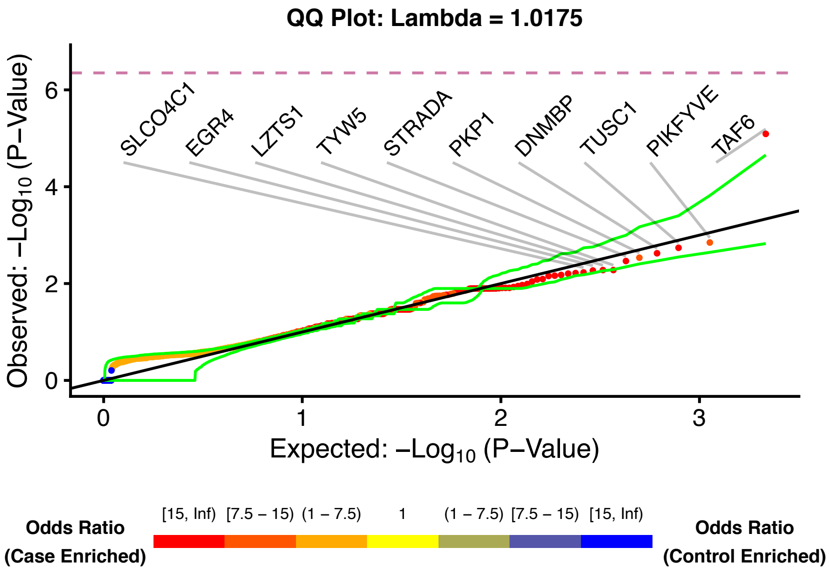

*eFigure 3: Ultra-Rare Synonymous Model Quantile-Quantile Plot for Unresolved Cohort*
The quantile-quantile plots for the protein-coding genes with at least one case or control carrier of an ultra-rare synonymous variant. All variants are ultra-rare (qualifying variants were defined as a minor allele frequency of less than 0.05% in internal case and control by cluster and absent in external reference cohorts). *P*-values were generated from the exact two-sided Cochran-Mantel-Haenszel (CMH) test by gene by cluster to indicate a different carrier status of cases in comparison to controls. Study-wide significance $p < 4.5 \times 10^{-7}$ after Bonferroni correction indicated by dashed line (see Collapsing by Gene and Statistical Enrichment. Top ten case enriched genes are labeled. Point coloring determined by CMH odds ratio. The green lines represent the 95% confidence interval.

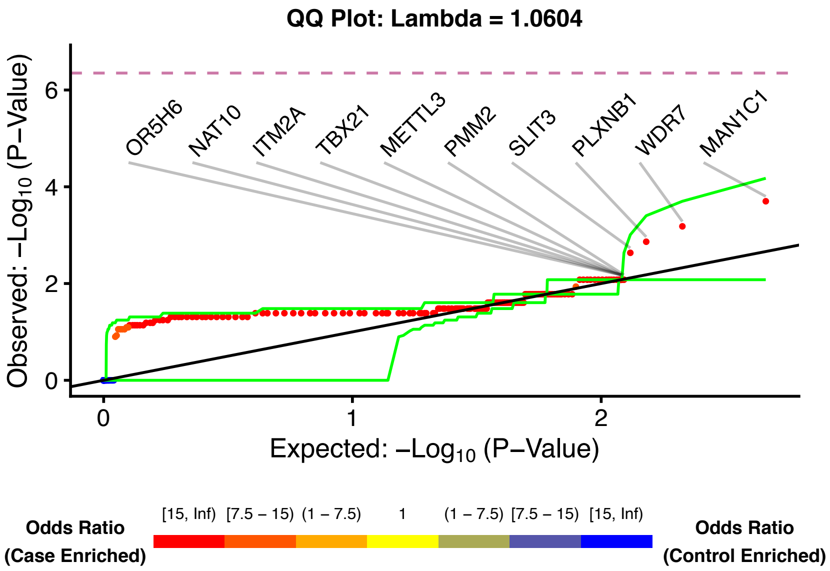

*eFigure 4: Ultra-Rare Synonymous Model Quantile-Quantile Plot for Respiratory Failure Cohort*

The quantile-quantile plots for the protein-coding genes with at least one case or control carrier of an ultra-rare synonymous variant. All variants are ultra-rare (qualifying variants were defined as a minor allele frequency of less than 0.05% in internal case and control by cluster and absent in external reference cohorts). P-values were generated from the exact two-sided Cochran-Mantel-Haenszel (CMH) test by gene by cluster to indicate a different carrier status of cases in comparison to controls. Study-wide significance $p < 4.5 \times 10^{-7}$ after Bonferroni correction indicated by dashed line (see Collapsing by Gene and Statistical Enrichment. Top ten case enriched genes are labeled. Point coloring determined by CMH odds ratio. The green lines represent the 95% confidence interval.

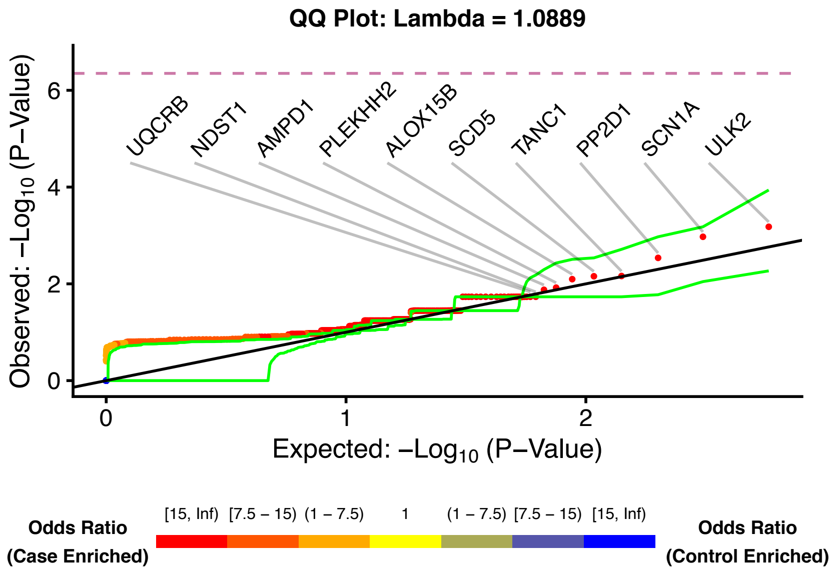

*eFigure 5: Ultra-Rare pLOF Model Quantile-Quantile Plot for Combined Cohort*

The quantile-quantile plots for the protein-coding genes with at least one case or control carrier of an ultra-rare pLOF variant. All variants are ultra-rare (qualifying variants were defined as a minor allele frequency of less than 0.05% in internal case and control by cluster and absent in external reference cohorts). P-values were generated from the exact two-sided Cochran-Mantel-Haenszel (CMH) test by gene by cluster to indicate a different carrier status of cases in comparison to controls. Study-wide significance $p < 4.5 \times 10^{-7}$ after Bonferroni correction indicated by dashed line (see Collapsing by Gene and Statistical Enrichment. Top ten case enriched genes are labeled. Point coloring determined by CMH odds ratio. The green lines represent the 95% confidence interval.

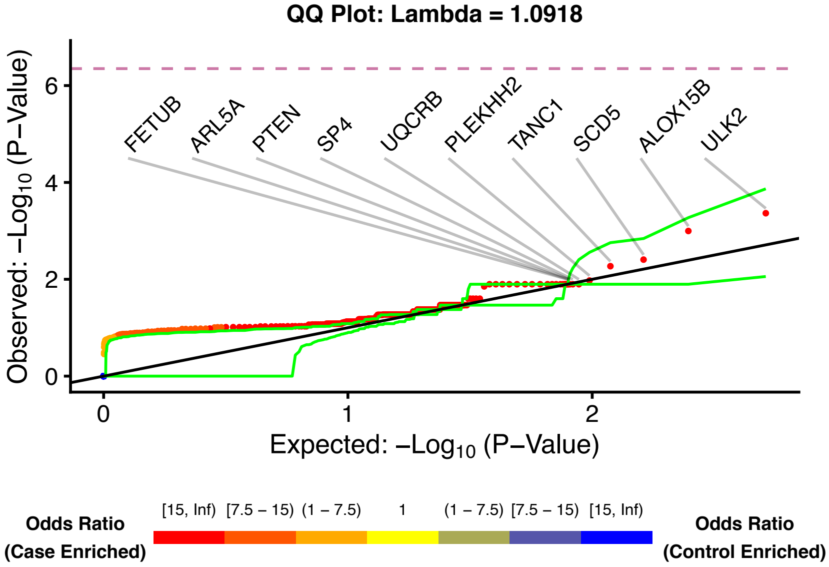

*eFigure 6: Ultra-Rare pLOF Model Quantile-Quantile Plot for Unresolved Cohort*

The quantile-quantile plots for the protein-coding genes with at least one case or control carrier of an ultra-rare pLOF variant. All variants are ultra-rare (qualifying variants were defined as a minor allele frequency of less than 0.05% in internal case and control by cluster and absent in external reference cohorts). P-values were generated from the exact two-sided Cochran-Mantel-Haenszel (CMH) test by gene by cluster to indicate a different carrier status of cases in comparison to controls. Study-wide significance $p < 4.5 \times 10^{-7}$ after Bonferroni correction indicated by dashed line (see Collapsing by Gene and Statistical Enrichment. Top ten case enriched genes are labeled. Point coloring determined by CMH odds ratio. The green lines represent the 95% confidence interval.

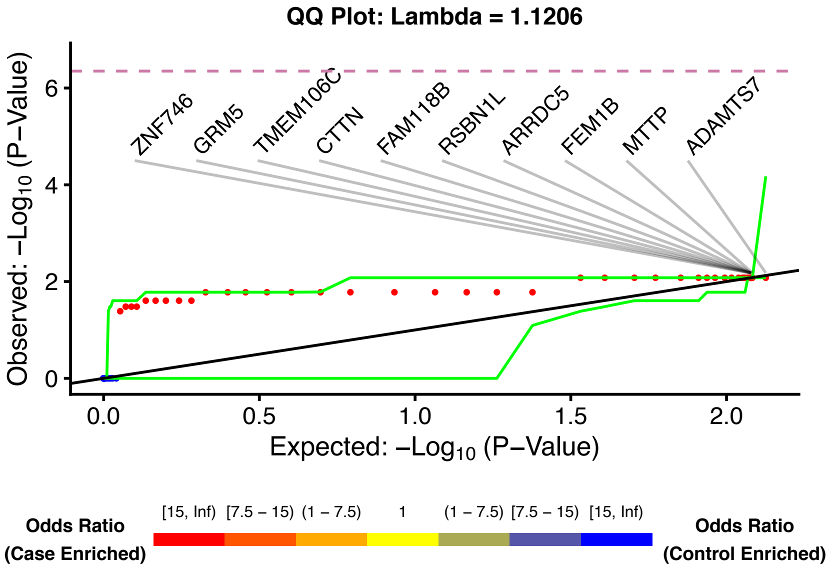

*eFigure 7: Ultra-Rare pLOF Model Quantile-Quantile Plot for Respiratory Failure*

The quantile-quantile plots for the protein-coding genes with at least one case or control carrier of an ultra-rare pLOF variant. All variants are ultra-rare (qualifying variants were defined as a minor allele frequency of less than 0.05% in internal case and control by cluster and absent in external reference cohorts). P-values were generated from the exact two-sided Cochran-Mantel-Haenszel (CMH) test by gene by cluster to indicate a different carrier status of cases in comparison to controls. Study-wide significance $p < 4.5 \times 10^{-7}$ after Bonferroni correction indicated by dashed line (see Collapsing by Gene and Statistical Enrichment. Top ten case enriched genes are labeled. Point coloring determined by CMH odds ratio. The green lines represent the 95% confidence interval.

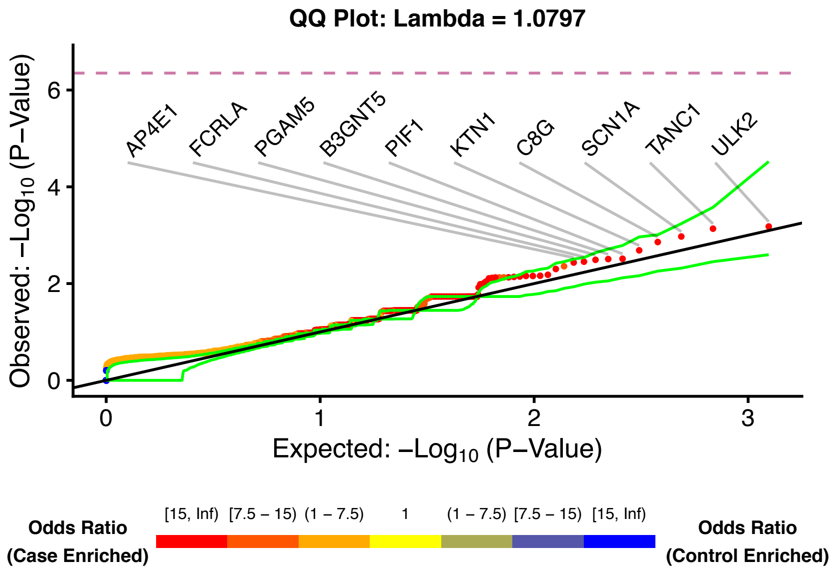

*eFigure 8: Flex pLOF Model Quantile-Quantile Plot for Combined*

The quantile-quantile plots for the protein-coding genes with at least one case or control carrier of a pLOF variant. All variants are ultra-rare (qualifying variants were defined as a minor allele frequency of less than 0.1% in external reference cohorts). P-values were generated from the exact two-sided Cochran-Mantel-Haenszel (CMH) test by gene by cluster to indicate a different carrier status of cases in comparison to controls. Study-wide significance $p < 4.5 \times 10^{-7}$ after Bonferroni correction indicated by dashed line (see Collapsing by Gene and Statistical Enrichment. Top ten case enriched genes are labeled. Point coloring determined by CMH odds ratio. The green lines represent the 95% confidence interval.

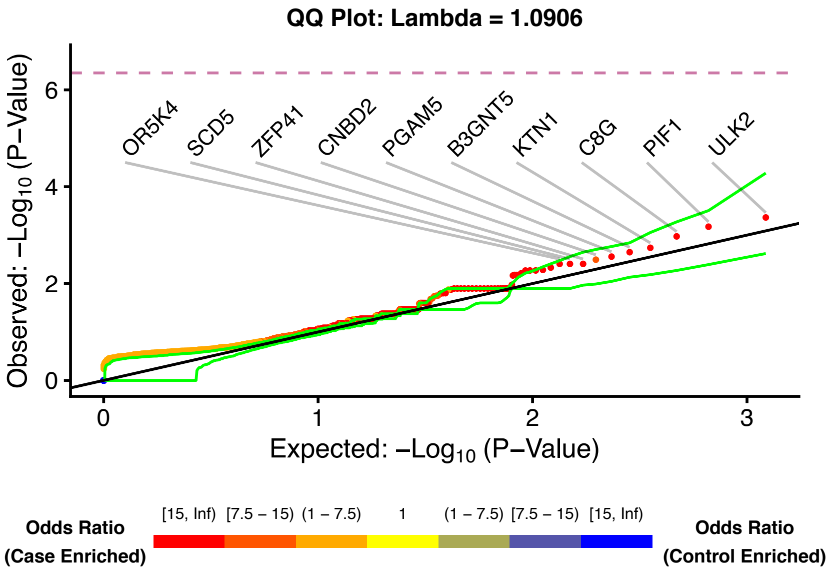

*eFigure 9: Flex pLOF Model Quantile-Quantile Plot for Unresolved Cohort*

The quantile-quantile plots for the protein-coding genes with at least one case or control carrier of a pLOF variant. All variants are ultra-rare (qualifying variants were defined as a minor allele frequency of less than 0.1% in external reference cohorts). P-values were generated from the exact two-sided Cochran-Mantel-Haenszel (CMH) test by gene by cluster to indicate a different carrier status of cases in comparison to controls. Study-wide significance $p < 4.5 \times 10^{-7}$ after Bonferroni correction indicated by dashed line (see Collapsing by Gene and Statistical Enrichment. Top ten case enriched genes are labeled. Point coloring determined by CMH odds ratio. The green lines represent the 95% confidence interval.

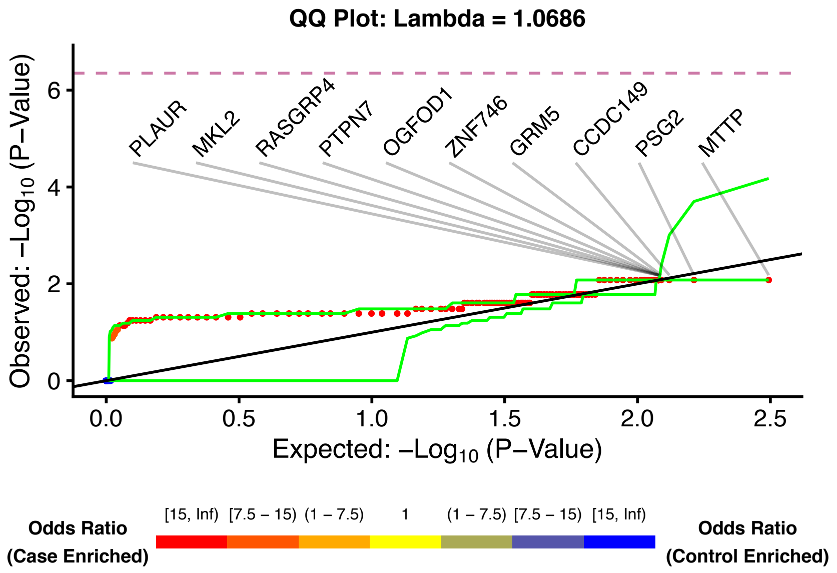

*eFigure 10: Flex pLOF Model Quantile-Quantile Plot for Respiratory Failure Cohort*
The quantile-quantile plots for the protein-coding genes with at least one case or control carrier of a pLOF variant. All variants are ultra-rare (qualifying variants were defined as a minor allele frequency of less than 0.1% in external reference cohorts). P-values were generated from the exact two-sided Cochran-Mantel-Haenszel (CMH) test by gene by cluster to indicate a different carrier status of cases in comparison to controls. Study-wide significance $p < 4.5 \times 10^{-7}$ after Bonferroni correction indicated by dashed line (see Collapsing by Gene and Statistical Enrichment. Top ten case enriched genes are labeled. Point coloring determined by CMH odds ratio. The green lines represent the 95% confidence interval.

**eMethods**

#### Study Design, Participants, and Consent

Children admitted to the PICU for trauma were excluded. Columbia University institutional review board approved protocols and informed consent was provided for the use of DNA in genetic research. Subjects enrolled at MSCH were referred by their CUIMC clinician and written informed consent was obtained through an institutional-review-board approved whole exome sequencing research protocols at the Institute for Genomic Medicine (IGM) (IRB# AAAO6702 and AAAO8410).[1] For subjects under 18 years old, informed written consent was provided by the child's legal guardian or the subject themselves when 18 years old or over.

#### Definition of Critical Illness

All probands were critically ill as defined by meeting any of the following criteria: (1) admission to the pediatric intensive care unit (PICU) at Morgan Stanley Children's Hospital of NewYork-Presbyterian (MSCH) – Columbia University Irving Medical Center (CUIMC) (n = 264), (2) PICU admission at another institution per the medical record (n = 1), (3) death (n = 18), or (4) respiratory failure on the general pediatric floor requiring continuous positive airway pressure (n = 2). Included probands from MSCH/CUIMC) were enrolled either during admission to the pediatric intensive care unit (PICU) or a subsequent visit. In addition, three children were recruited from the general pediatric floor.

#### Respiratory Failure Cohort

The "respiratory failure cohort" was composed of probands from MSCH and OCME. MSCH probands were included if they had no contributing medical history of other than asthma o reactive airway disease, gestational age greater than or equal to 36 weeks (one proband did not have a gestational age included in the chart), and a common viral illness noted in the medical chart (eTable 2), and required invasive or non-invasive positive pressure ventilation (eTable 2). OCME probands were included if the limited medical history excluded any indication of chronic disease (eTable 1).

#### Controls

The same control samples were used as a base of comparison for each of the three case cohorts.

#### Phenotyping Children with Critical Illness

To qualitatively describe the heterogenous phenotypes present in the combined cohort, we mapped PICU admission notes (or general pediatric admission notes when no PICU note was available) to the Human Phenotype Ontology (eTable 6).[2,3] We used doc2hpo for an initial conversion to HPO concept IDs and then further manually reviewed and edited the mapped concepts.[4] For the OCME cohort, limited clinical information was used (eTable 1).

#### WES and WGS Data Generation

Exomes were captured with multiple capture kits and sequenced according to standard protocols on Illumina's HiSeq 2000, HiSeq 2500, and NovaSeq 6000 (Illumina, San Diego, CA, USA) platform with 150 bp paired-end reads. Genomes were sequenced according to standard protocols on Illumina's NovaSeq 6000 (Illumina, San Diego, CA, USA) platform. Possible bias introduced by using WES from different capture kits (eTable 22) and WGS in collapsing is corrected during coverage harmonization (see Collapsing Coverage Harmonization).[5,6]

#### Diagnostic Analysis

For diagnostic analysis, sample preparation, alignment and variant calling followed the same pipeline as in collapsing analysis (see WES and WGS Data Generation and Alignment and Variant Calling). WES data were analyzed using the qualifying genotype approach as previously been described previously. A combination of the American College of Medical Genetics and Association for Molecular Pathology variant classification as well as clinical context and clinical correlation from phenotype experts/referring providers was used to assess variant causality.[1,7-10] The OCME cohort was reviewed as well although limited clinical data made a full diagnostic analysis impossible. Probands were deemed resolve if their phenotype was fully or partially explained by either the research exome or any other clinical genetics data available by chart review. The summary of these data can found in eTable 5.

#### Alignment and Variant Calling

Both affected individuals and control individuals were processed with the same IGM bioinformatic pipeline for variant calling. Reads were aligned to human reference GRCh37 via DRAGEN (Edico Genome, San Diego, CA, USA)[11] and duplicates were marked with Picard (Broad Institute, Boston, MA, USA). Variants were called according to the Genome Analysis Toolkit (GATK - Broad Institute, Boston, MA, USA) Best Practices recommendations v.3.6.[12,13] Finally, variants were annotated with ClinEff.[14] Custom annotations including Genome Aggregation Database (gnomAD) v.2.1 frequencies[15], Exome Aggregation Consortium (ExAC)[16] frequencies, and loss-of-function observed/expected upper bound fraction (LOEUF)[15] scores and deciles were added via the IGM's in-house analysis tool for annotated variants (ATAV) platform.[17]

#### Collapsing Analysis Overview

The collapsing analysis implemented in this manuscript[5] takes the following steps: i) high-quality samples and selected controls with non-overlapping phenotypes are chosen and related individuals are removed (see Analyzed Cohort Definitions, Control Samples, and Collapsing Sample Quality Control), ii) match cases and controls in clusters based on geographic ancestry (see Collapsing Geographic Ancestry Clustering), iii) perform coverage harmonization separately in each cluster (see Collapsing Coverage Harmonization), iv) create models with specific QV criteria (see Table S3, Collapsing Variant Quality Control and Collapsing Model Specification), v) collapse by gene or gene set and assign indicator variable (0/1) to each case and control based on the absence/present of a QV in the gene or gene set, and vi) test for association between case/control status and indicator variable (see Collapsing by Gene and Statistical Enrichment and Collapsing Gene-set Enrichment Testing), and vi) visualize results (see Collapsing Quantile-Quantile (QQ) Plots and Genomic Inflation Factor).

#### Collapsing Sample Quality Control

The same quality control standards were applied to cases and controls. We included only samples with at least 90% of the consensus coding sequence (CCDS release 20)[18] covered at a minimum of 10x, less or equal 2% contamination levels according to VerifyBamID[19], and single nucleotide variants (SNVs) and indels overlapping the Single Nucleotide Polymorphism database (dbSNP)[20] at least 85% and 80%, respectively. We excluded with a discordance between self-declared and sequence-derived gender to prevent phenotype-genotype mismatch. We utilized KING to detect related individuals and removed one of each pair that had an inferred relationship of second-degree or closer while favoring the inclusion of cases over controls and well-covered over poorly-covered.[21] Of the 285 samples in the combined cohort, three samples from the MSCH cohort were excluded due to low quality. No OCME samples were excluded.

#### Collapsing Geographic Ancestry Clustering

It is important to correct for the underlying rate of variation in samples of different geographic ancestry in case/control experimental designs.[22-24] Clusters containing at least 20 cases and 20 controls were used for all analyses based on collapsing clusters (Figures 1, 2, and 3) [6] All clusters underwent coverage harmonization (see Collapsing coverage harmonization in Methods). A pre-trained neural-network generated probability estimates for each of six groups (European, African, Latino, East Asian, South Asian and Middle Eastern). A geographic ancestry label was assigned to each sample using a 95% probability cut-off. “Admixed” samples were those that did not reach 95% for any of the ancestry groups (eFigure 1). The following steps were performed separately for each of the three analyzed case-control cohorts (see Analyzed Cohort Definitions). To check the quality of the clusters, we performed further dimensionality reduction using the Uniform Manifold Approximation and Projection (UMAP)[25] on the first six PCs to disentangle subcontinental structure, which is then reflected in the cluster membership.[26,27] (1) The combined and unresolved cohorts included multiple clusters and used the Cochran-Mantel-Haenszel test for statistical association testing. The respiratory failure cohort included only one cluster and used Fisher's exact test for statistical association testing.

#### Collapsing Coverage Harmonization

Coverage differences between cases and controls introduce bias because variants can only be called with sufficient coverage. As described previously[5,6,28-30], to reduce the influence of coverage differences caused by different capture kits, inclusion of both WES and WGS or sequencing depth in general, we used a site-based pruning approach and removed sites where the absolute difference in percentages of cases compared to controls with at least 10x coverage was greater than 7.0%. We performed coverage harmonization on each cluster independently (see Collapsing Geographic Ancestry Clustering). This resulted in three sets coverage maps (eFigure 1).

#### Collapsing Variant Quality Control

In each cluster, we called variants at bases available for variant calling per cluster-specific coverage harmonization (see Collapsing Coverage Harmonization). Only variants meeting the following qualifications were considered for analysis: i) at least 10x coverage of the site, ii) quality score (QUAL) $\geq$ 50, iii) genotype quality score (GQ) $\geq$ 20, iv) quality by depth score (QD)

≥ 5, v) mapping quality score (MQ) ≥ 40, vi) read position rank sum score (RPRS) ≥ -3, vii) mapping quality rank sum score (MQRS) ≥ -10, viii) Fisher's strand bias score (FS) ≤ 60 (SNVs) or ≤ 200 (indels), ix) strand odds ratio (SOR) ≤ 3 (SNVs) or ≤ 10 (indels), x) GATK Variant Quality Score Recalibration filter "PASS", xi) alternate allele fraction for heterozygous calls ≥ 0.3, xii) within the CCDS inclusive of two base intronic extensions to accommodate canonical splice variants, xiii) a proportion expression across transcripts (pext) value (when available) greater than or equal to 1/10 the maximum pext value for that gene were removed as they are unlikely to affect translated mRNA[31], and xiv) located outside regions with highly repetitive elements to reduce false-positivity.[32] Sequencing artifacts as described previously[28] and low quality variants per Exome Aggregation Consortium[33], gnomAD[15], or the Exome Variant Server were excluded (see Web Resources). All predicted loss-of-function (pLOF) variants (stop gain, frameshift, splice acceptor, and splice donor variants) were filtered with Loss-Of-Function Transcript Effect Estimator (LOFTEE) to remove likely false-positive pLOFs.[15]

#### Collapsing Model Specification

Each collapsing model depends on the definition of a qualifying variant (QV) with parameters designed to enriched for real variant calls with strong functional effects (eTable 3).[5,34] Model parameters included external minor allele frequency (MAF), internal allele frequency, variant effect, and *in-silico filters*. External frequency filters in gnomAD and ExAC which could be either "ultra-rare" (absent) or "flex" (MAF < 0.1%). For the flex model, MAF was filtered at a population specific level. For ExAC, populations included afr, amr, nfe, fin, eas, sas. For gnomAD exomes, populations included afr, amr, asj, eas, sas, fin, and nfe. For gnomAD genomes, the MAF filter was applied to the full population. For the purposes of gene-set analysis (Figures 2 and 3), a further subset "rare but public" was defined which removed all ultra-rare variants leaving only variants that are rare but still present in gnomAD or ExAC. Internal allele frequencies were applied by cluster. For ultra-rare models, variants were excluded with an internal allele frequency greater than 0.05% applied to the combined case-control call set by cluster excluding one allele to allow for clusters in which one allele might exceed that allele frequency threshold.[31] For flex models, the internal allele frequency filter was set at 0.1%.

#### Collapsing by Gene and Statistical Enrichment

From the collapsing matrices of each cluster, we extracted the number of cases/controls with and without a QV per gene and used the exact two-sided Cochran-Mantel-Haenszel (CMH) test to test for an enrichment of qualifying variants in the case or control group (eTable 2) while controlling for cluster membership.[6,29,35,36] (2) The respiratory failure cohort included only one cluster large enough for analysis. For models analyzed for this cohort, an individual-by-gene matrix was created as above for the single cluster. We implemented a two-tailed Fisher's exact test to identify genes where there was a significant enrichment of qualifying variants in the case or control group.[37]

To visualize our results and ensure appropriate genomic inflation, we created quantile-quantile (QQ) plots (described below). The synonymous model was used as a putatively negative control for each cohort (eFigures 2 - 4, eTables 8 – 10). We defined a study-wide Bonferroni multiplicity-adjusted significance threshold of $p < 4.5 \times 10^{-7}$ (0.05 / [18650 CCDS genes × 6 non-synonymous models]). Model details for the six non-synonymous models can be found in eTable 3. The top 50 ranked genes for all nine models can be found in the supplemental tables (eTables 8 - 16). The membership of each gene in the following gene-sets is also indicated: (O) disease association in the Online Mendelian Inheritance in Man as of 02/16/2021 (OMIM, see Web Resources) (see Gene-Set Enrichment Testing).

#### Collapsing Quantile-Quantile (QQ) Plots and Genomic Inflation Factor λ

For each model, we plotted expected vs. observed *p*-values for our collapsing by gene enrichment results. We generated empirical (permutation-based) expected probability distributions using one of two methods for each model independently. (1) For models using the combined and unresolved cohort which used multiple clusters, we used a process previously described.[6,29] For each cluster, the original case and control labels were randomly permuted while the rest of the gene by sample matrix was kept fixed. For each cluster we extracted the number of newly labeled cases/controls with and without a QV per gene and used the CMH test to test for an association between case/control status and QV status while controlling for cluster membership (see Collapsing by Gene and Statistical Enrichment). This process was repeated 1,000 times to create an empirical distribution of 1,000 *p*-values for each gene, and for each permutation the *p*-values were ordered. (2) For the respiratory failure cohort in which only one cluster was used, we randomly permutated the case/control labels in the single cluster.[5,28,38,39] After each permutation, A two-tailed Fisher's exact test was performed to test for an association between case/control status and QV status (see Collapsing by gene and statistical enrichment). This process was repeated 1,000 times to create an empirical distribution of 1,000 *p*-values for each gene and for each permutation, the *p*-values were ordered.

Empirical estimates of the expected ordered *p*-values were represented by the mean of each rank-ordered estimate across the 1,000 permutations (i.e., the average 1st order statistic, the average 2nd order statistic, etc.). The negative logarithms of the expected and observed *p*-values were plotted to get permutation-based QQ plots. We estimated the genomic inflation factor λ based on the permutation-based expected *p*-values using a regression method as described previously.[28,37]

#### Collapsing Gene-Set Enrichment Testing

Biologically informed gene-sets can reveal important pathways or gene characteristics by aggregated signal across related genes.[5,37] Association between case/control status and harboring a variant in a gene-set was tested in two ways. (1) For the combined and unresolved cohorts which included multiple clusters, we extracted the number of cases/controls with and without at least one QV among any of the genes in each of the gene-sets and used the exact two-sided CMH test[6,35,36] to test for an association between case/control status and QV status while controlling for cluster membership. (2) For the respiratory failure cohort which included only one cluster, we extracted the number of cases/controls with and without at least one QV among any of the genes in each of the gene-sets and used a two-tailed Fisher's exact test to test for an association between case/control status and QV status.[37]

We used a false discovery rate (FDR) correction for multiple comparisons. We performed 20 CMH tests or FETs to determine odds ratios for gene-set enrichment testing and defined a significant enrichment at FDR < 0.05.

#### Unbiased pLOF Enrichment Analysis

Given the rarity of pediatric critical illness, we hypothesized that the combined cohort would be more likely to harbor pLOF variants than controls and that these variants would exist in genes "intolerant" to variation (i.e., genes in which few pLOF variants are found in otherwise healthy individuals).[15,40] The empirical *p*-values of the 1,860 tests were determined by permutation. We randomly shuffled the case/control labels within each cluster and re-calculated the *p*-value. This was performed 100,000 times leaving 100,000 permuted *p*-values for each of the 1,860 gene-sets. The empirically derived *p*-value at each gene-set was then determined by the fraction of permuted *p*-values less than the actual *p*-value for that gene-set. We identified the LOEUF value with the most significant threshold and used the gene-set defined by this LOEUF threshold for forest plots (see Collapsing Gene-Set Enrichment Testing).

#### *De Novo* Mutation Calling, Filtering and Analysis

We used ATAV's "—list-trio" function (ATAV v7.2.1) to screen for *de novo* variants. The initial set of candidate *de novo* variants met the following quality control thresholds: i) AD Alt ≥3, ii) QC Filter Pass, iii) QUAL ≥ 50, iv) GQ ≥ 20, v) MQ ≥ 40, vi) variant site covered in both parents with at least 10 reads, vii) variant absent in parents, viii) variant absent in gnomAD exomes and genomes, ix) absent in IGM controls, and x) child het carrier ≥ 10% alt read OR child hom carrier ≥ 80% alt read. To further improve the quality of *de novo* calls, we imposed additional criteria: i) variant located outside regions with highly repetitive elements to reduce false-positivity,[32] and ii) SNV VQSR tranche < 99. Finally, all loss-of-function *de novo* variants were visually inspected in IGV to confirm the underlying alignment. Any variant call that failed visual inspection was excluded.

Using denovolyzeR, synonymous, missense and loss-of-function are analyzed in addition to combining missense and loss-of-function to capture protein-altering variants.[41-43], The tool allows for analyses of all genes or gene-sets. We focused on genes without a disease association in OMIM. Loss-of-function variants were those with frameshift, splice_acceptor_variant, splice_donor_variant, and stop_gained effects.

#### Determination of Disease-Gene Association

Gene-disease associations were determined from the Online Mendelian Inheritance in Man (OMIM, see web resources) with the following methods.[44]. genemap2.txt was downloaded on 2/16/2021. Genes were filtered to remove those with blank phenotypes or phenotypes that begin with "?" (indicating a relationship between the phenotype and gene is provisional) or "[" (indicating a non-disease phenotype). The remaining genes are considered to be associated with disease. Using this method, 3,964 genes were determined to have a disease association in OMIM. The remaining genes were our deemed to be without a disease association. The "Primary Immunodeficiency" gene set was 424 genes drawn from the Invitae Primary Immunodeficiency Panel (Test code: 08100, https://www.invitae.com/en/providers/test-catalog/test-08100). The Viral Immunodeficiency gene set was 13 genes drawn from Zhang et al.[45]

#### Data Analysis and Display

Unless otherwise noted in the methods, data analysis and visualization were performed with R (v.3.6.0).[46]

#### Web Resources

ATAV, https://github.com/igm-team/atav
Consensus Coding Sequence, https://www.ncbi.nlm.nih.gov/CCDS/
Exome Aggregation Consortium (ExAC), http://exac.broadinstitute.org
Exome Variant Server, https://evs.gs.washington.edu/EVS/
Genome Aggregation Database (gnomAD), https://gnomad.broadinstitute.org
Genome Analysis Toolkit (GATK), https://gatk.broadinstitute.org/hc/en-us
NIH Genomic Data Sharing Policy, https://osp.od.nih.gov/scientific-sharing/policies/

OMIM, https://www.omim.org
Picard, http://broadinstitute.github.io/picard/
R, https://www.R-project.org/
